# OpenPrescribing Hospitals: An open access analytics platform for hospital medicines use in England

**DOI:** 10.1101/2025.11.13.25340060

**Authors:** Louis Fisher, Christopher Wood, Andrew Brown, Richard Croker, Stephen Black, Helen J Curtis, Rose Higgins, Arina Tamborska, Sebastian Bacon, Ben Goldacre, Brian MacKenna, Victoria Speed

**Affiliations:** Bennett Institute for Applied Data Science, Nuffield Department of Primary Care Health Sciences, University of Oxford, OX2 6GG, UK; King’s Thrombosis Centre, King’s College Hospital, Denmark Hill, London, SE5 9RS

## Abstract

Primary care prescribing data for general practices in England has been openly published since 2011. The OpenPrescribing platform provides an open, transparent and easy-to-use interface to this data. OpenPrescribing is widely used by NHS professionals, the public and by industry and has supported analyses to identify cost-saving opportunities and unsafe prescribing in primary care in England. The Secondary Care Medicines Data, published since July 2021 makes analogous data for medicines issued in NHS trusts in England available for the first time. As with primary care prescribing data, this dataset requires extensive domain and technical knowledge to access, clean, transform and link to other external sources before it can be used effectively. We have developed the OpenPrescribing Hospitals platform to provide an intuitive interface to the dataset, built on top of a robust data pipeline that removes the barriers to the dataset’s use. This platform is openly accessible, with transparent methodology and aims to deliver the same benefits as OpenPrescribing. Here we describe why the platform is needed, its key features, and highlight potential use cases. We encourage adoption of the platform by clinicians, academics and policy advisors to support rapid local feedback against national medicines standards and to monitor the extent and variation of uptake of new medicines.

## Introduction

Data on prescribing in NHS-funded general practices has been openly available since 2011^1^. The scrutiny of prescribing data is embedded into routine care using both open and restricted access analytic platforms^2–4^. These tools are commonly used by medicines optimisation teams who provide feedback to GPs to action better value, safer and more environmentally friendly prescribing. Until recently, analogous data for medicines usage in secondary care in England was not openly available.

In July 2020, we advocated for this previously inaccessible data to be made openly available^5^. In July 2021 it was made publicly available for the first time in the form of the Secondary Care Medicines Data (SCMD)^6^. This dataset provides detailed information on the usage of individual medicines at the level of individual NHS trusts by capturing stock issuing data from hospital pharmacies in all NHS acute, teaching, specialist, mental health and community trusts in England^6^. The dataset is collated by Rx-Info, a UK-based healthcare analytics company that provides medicines data and analytics software to the NHS^7^. It is hosted and published on behalf of NHS England via the NHS Business Services Authority (BSA) Open Data Portal^8^.

Since its publication, studies carried out using the SCMD showcase the benefits of making this data openly available and its potential for future use^9–13^. However, this rich data source remains underutilised. As with other NHS datasets, there are barriers to its use^14^: multiple versions of the data exist^6,15^; monthly data is made available as separate files which must be downloaded, aggregated and processed; and linkage to external data sources is required to contextualise the data (e.g. geographical location of organisations from the Organisation Data Service^16^). A small number of dashboards exist, which attempt to handle this complexity and provide high level insights for NHS trusts^17,18^. Platforms offering richer datasets and more advanced analytical capabilities, are typically restricted to NHS users^19–22^. This limited access constrains broader efforts to benchmark activity; explore measures of value, safety, and evidence-based practice; and share examples of best practice more widely.

Since 2015, we have provided the OpenPrescribing platform^23,24^, which supports a wide range of analyses of primary care prescribing data. Building on this experience, we have developed OpenPrescribing Hospitals. We aim to achieve the same safety, efficiency and innovation benefits from the SCMD that OpenPrescribing has already demonstrated in primary care^25–28^. The platform is openly available at https://hospitals.openprescribing.net, free to use, and completely transparent. Here, we describe the features of the platform that support exploration of the SCMD, and provide examples of how these features can be used to understand medicines usage and inform clinical practice.

## Methods

### Implementation

OpenPrescribing Hospitals (https://hospitals.openprescribing.net) is a web application that provides analytics and insights into NHS hospital medicines data.

The data available through the OpenPrescribing Hospitals platform is a curated aggregation of multiple data sources (**Box 1**). The primary data source is the SCMD^6^.

## Box 1. Data sources used by OpenPrescribing Hospitals

**Secondary Care Medicines Dataset (SCMD)** - the primary data source, containing information about medicines issued in hospitals. This data is published by the NHS BSA and is available to download from the NHS BSA SCMD data download page^15^ or using the NHSBSA Open Data Portal application programming interface (API)^29^. It is also accessible through ePACT2 for registered users^30^.

**Dictionary of Medicines and Devices (dm+d)** - a standardised dictionary of medicines and devices, providing supplementary information about products included in the SCMD. For example, information about the ingredients within a product and its route of administration. The dm+d is updated weekly and published by the NHS BSA^31^.

**Organisational Data Service (ODS)** - individual organisations in the SCMD are identified by their ODS^16^ code. Basic information about these organisations, such as their name, location and relationship to other organisations can be obtained using the ODS Data Search And Export tool^32^ or NHS ODS Organisation Reference Data API^33^.

**Anatomical Therapeutic Chemical/Defined Daily Dose (ATC/DDD)** - The Anatomical Therapeutic Chemical (ATC) system provides a classification for medicinal substances based on their main therapeutic, pharmacological, and chemical properties. In combination with the Defined Daily Dose (DDD)—a unit that enables comparison of medicines consumption—the ATC system supports nuanced analysis of usage across groups of medicines. The ATC/DDD index is maintained by The World Health Organisation (WHO) Collaborating Centre for Drug Statistics Methodology^34^ and published openly^35^. A machine readable version is available at a cost.

All data sources outlined in **Box 1** are dynamic and routinely updated. To support ongoing updates, we have developed a data pipeline to import, transform, validate and link these sources as part of the OpenPrescribing Hospitals platform. This pipeline is managed using the workflow orchestration tool Prefect^36^. The pipeline includes:

● Linkage of the SCMD data to the dm+d to identify further product information, such as the ingredients they contain, or their routes of administration.
● Linkage of the SCMD to the ODS to identify NHS trust information, including their relationships to other NHS organisations such as the Integrated Care Board (ICB) they belong to, or other NHS trusts with which they have merged or been acquired by. This supports aggregation of data for historical trusts into their successor trust in the features described below.
● Linkage of the SCMD to ATC/DDD and dm+d to enable calculation of alternative measures of quantity (**Box 2**).

## Box 2. OpenPrescribing Hospitals quantity metrics

**SCMD quantity** - the quantity as reported in the SCMD with the following adjustments:

● Historical dm+d codes used to identify products are mapped to their most recent dm+d code so that the data for a product identified using multiple identifiers in the raw data is aggregated under a single code.
● Reported quantities are converted to quantities in basis units to enable easier aggregation and comparison (e.g. quantities in *micrograms* are converted to quantities in *grams*, *microlitres* to *millilitres*).

Every product has an *SCMD quantity*, so this is a reliable way to look at the issuing of single products. However, it uses a wide range of units of measure (e.g. tablets, sachets and millilitres) which limits its use for comparing multiple products.

**Unit dose quantity** - a *unit dose* is a single measured quantity of a medicine, packaged for administration to a patient in a single unit. This is different to *dose*, which is the quantity of medication given as part of a broader dosage regimen that includes frequency and duration. Unit doses are only defined for products with a *discrete* unit dose form in the dm+d.

**Ingredient quantity** - the amount of the active ingredients within a product. Ingredient quantity is calculated for all specified ingredients within a product where the unit of measurement for the ingredient is compatible with the reported *SCMD quantity*.

**Defined Daily Dose quantity** - a DDD is a standardised unit of drug utilisation, defined by the WHO, that allows comparison of drug use between different countries, regions and healthcare settings. DDDs are only calculated for products which have been mapped to an ATC code and which have a compatible unit of measure with the specified DDD for that ATC code.

Data is stored and processed using Google BigQuery^37^. This data is then used to populate a PostgreSQL^38^ database, optimised to efficiently handle the various ways users interact with the data through the OpenPrescribing Hospitals web application. For example: supporting a user searching across all products reported in the SCMD by name, product code, ingredients they contain, or their therapeutic use; or querying the data for information on the quantity of a set of medicines issued over time in a subset of NHS trusts.

The web application is implemented using Django^39^, Svelte^40^ and TailwindCSS^41^. Charts are generated using the Highcharts library^42^. The platform is deployed using DigitalOcean App Platform^43^. Web analytics are measured using Plausible Analytics^44^. All code for the data pipeline and platform is openly available on GitHub^45^.

### Operation

The OpenPrescribing Hospitals platform has three primary features: *Submission History*, *Measures* and *Analyse* which are described in more detail below. Documentation and answers to frequently asked questions are available on the platform^46^.

#### Submission History

The SCMD contains processed pharmacy stock control data submitted by individual hospital trusts. This presents several challenges:

● Data submitted by individual trusts can be incomplete or inconsistent^47,48^.
● NHS organisations change over time as trusts open, close, merge or are acquired^49^.
● Historical stock control data can be amended after submission. As a result, data in the SCMD is first made available as provisional data before it is finalised annually^50^.

These challenges have to be considered when interpreting charts generated from the SCMD using the OpenPrescribing Hospitals platform. The *Submission History* feature aims to support users to navigate these complications by showing:

● All of the NHS trusts included in the SCMD.
● Data submission completeness and consistency over time.
● Organisational changes such as mergers and acquisitions between trusts.
● Whether the data each month represents provisional or finalised data.

The *Submission History* feature is demonstrated in **Figure 1**. It displays the monthly number of unique products each NHS trust submits to the SCMD. Months where trusts submit no data are represented by red bars. The height of each blue monthly bar represents the number of unique products reported within the SCMD for each trust that month.

**Figure 1.**
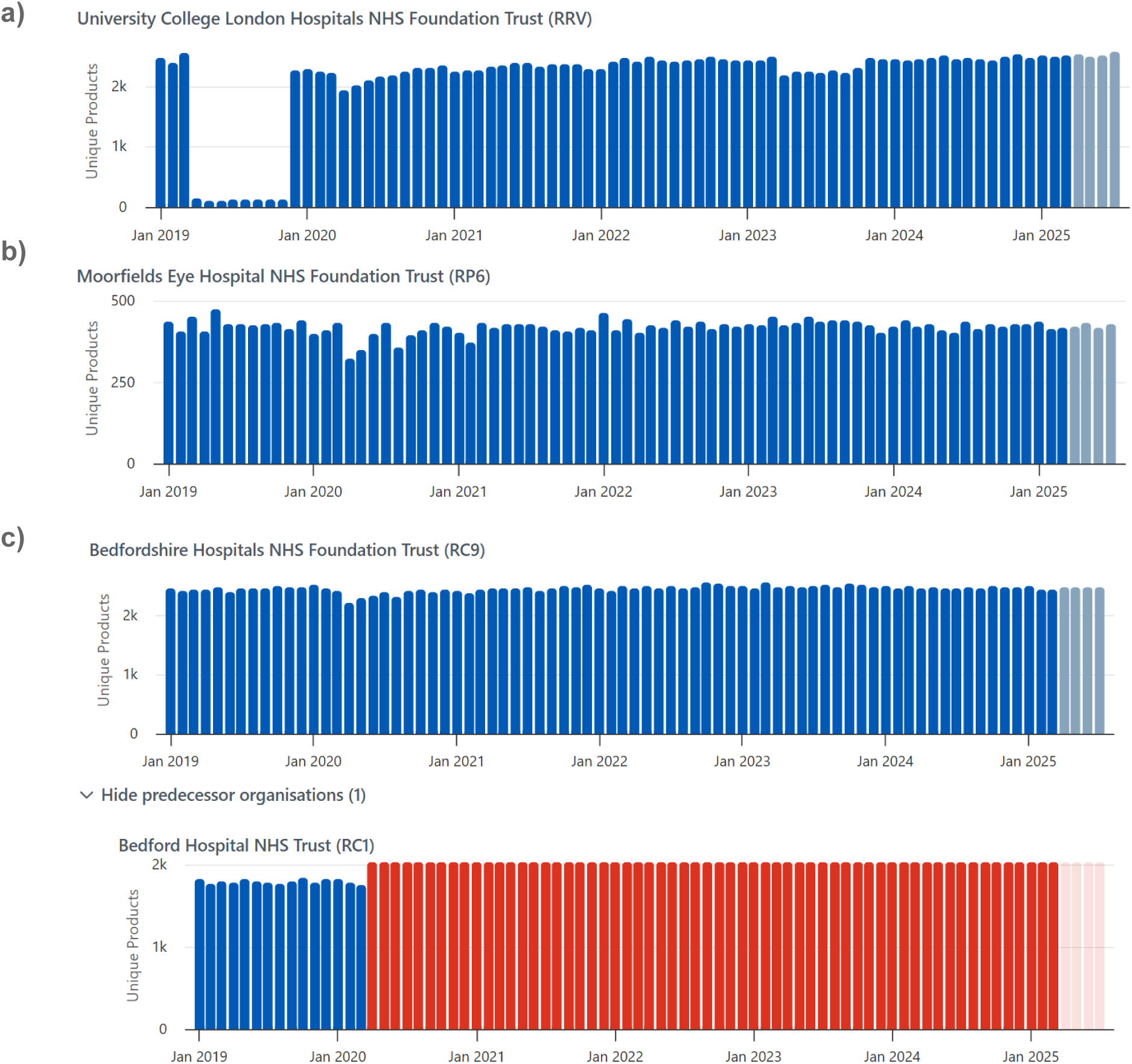
Submission history for SCMD as displayed on OpenPrescribing Hospitals for **a)** University College London Hospitals NHS Foundation Trust, which shows inconsistent number of products included in monthly submissions **b)** Moorfields Eye Hospital NHS Foundation Trust, a trust with a clinical speciality, which is reflected in the lower number of unique products the trust issues **c)** Bedfordshire Hospitals NHS Foundation Trust and its predecessor organisation, Bedford Hospital NHS Trust, which shows no submissions from April 2020. Red bars represent months where no products were reported in the SCMD for that trust. Transparent bars represent months where the SCMD is published as provisional data awaiting finalisation

For some trusts, this can indicate periods where the set of products they submit data for is incomplete (**Figure 1a**). Some variation in the number of unique products reported each month is expected, reflecting trust size, degree of specialisation and differences in the local formulary. For example, smaller more specialised trusts are more likely to issue a smaller number of products (eg. Moorfields Eye Hospital, **Figure 1b**). Where trusts have merged or been acquired, the *Submission History* groups these trusts with their successor **(Figure 1c)**.

In this example, the number of products for the successor trust (Bedfordshire Hospitals NHS Foundation Trust), represents the number of unique products issued in that trust and its predecessor (Bedford Hospital NHS Trust). Finally, this feature shows the status of the SCMD each month. Solid coloured red/blue bars represent finalised data and lighter coloured red/blue bars represent months where the data remains provisional.

#### Measures

*Measures* quantify activity for a clinical area of interest. They can be used to drive targeted quality improvement work by highlighting variation, and identifying potential areas for improvement in evidence-based practice, prescribing safety, and cost-effectiveness. *Measures* are based on national medicines recommendations, for example, NHS England medicines optimisation opportunities^51^ or prescribing guidance within the British National Formulary^52^.

For each measure on OpenPrescribing Hospitals, we describe:

**● What is this measure?** A brief description of what the measure captures.
**● How is it calculated?** An explanation of how products in the measure are chosen, what measure of quantity is used and how the measure is calculated from these quantities.
**● Why does it matter?** An indication of why the measure is important to monitor. For example, alignment with commissioning recommendations.
**● Which trusts are included?** Trusts are only included in a given measure if they issue the products included in the measure. This gives an indication of the number of trusts included.

The supporting text aims to make all measures easily understandable, transparent and reproducible to a general audience.

Measures are visualised on a time series chart with four modes: *NHS Trust, ICB, Region and National*. In *NHS Trust* mode, a percentile chart is displayed, on which individual trusts can be selected and displayed for comparison. **Figure 2** shows the charts available for a measure looking at the uptake of Phesgo (a new breast cancer treatment). In **Figure 2a**, the percentiles chart is presented with data for two hospital trusts overlaid. This mode is designed to clearly show the extent of variation across NHS trusts and how an individual trust compares to their peers. Below the time series chart, we show the products included in the measure, and their unit of measure. (**Figure 2b**).

**Figure 2.**
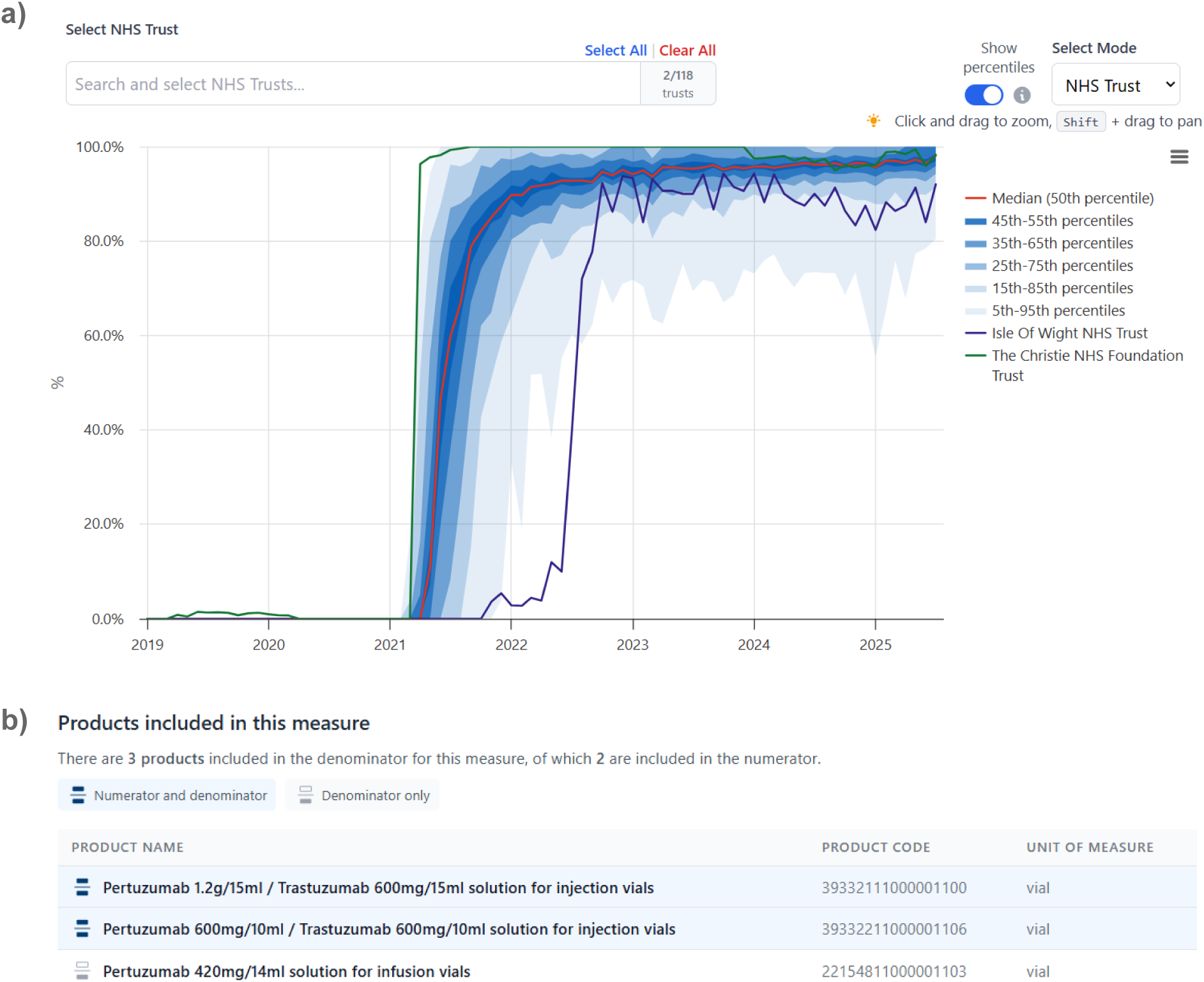
An example measure from OpenPrescribing Hospitals in NHS Trust mode showing **a)** time series chart with percentiles and individual trusts selected **b)** products included in the numerator and denominator of the measure.

### Analyse

The *Analyse* feature supports user-defined analyses against the underlying data. Analyses are specified using the *Analysis builder*, which supports:

● Free text search and selection of products of interest (**Figure 3a**). Products can be searched using:

○ **Product name or identifier codes** - products can be selected individually, or as groups of related products. For example, all strengths of codeine (dm+d Virtual Therapeutic Moiety code (VTM) 775354007) (e.g. codeine 30mg tablets, codeine 60mg tablets) can be selected.
○ **Ingredient names** - selecting an ingredient selects all products containing that ingredient. For example, using *Ingredient*, users could select all products containing paracetamol, including combination products such as co-codamol (paracetamol and codeine).
○ **ATC name or code** - selecting an ATC level will select all products matched to that ATC level or any of its children. For example, a user could select all ‘proton pump inhibitors’ (ATC code: A02BC).
● Optional free text search and selection of NHS trusts of interest. All trusts reporting data in the SCMD are included, with predecessor trusts shown with their successor. Predecessor trusts are not selectable individually.
● Selection of the quantity type (**Box 2**) to use for the analysis, using the advanced options. The most appropriate quantity type for an analysis is automatically decided based on the products selected (**Figure S1**), but can be manually specified.

**Figure 3.**
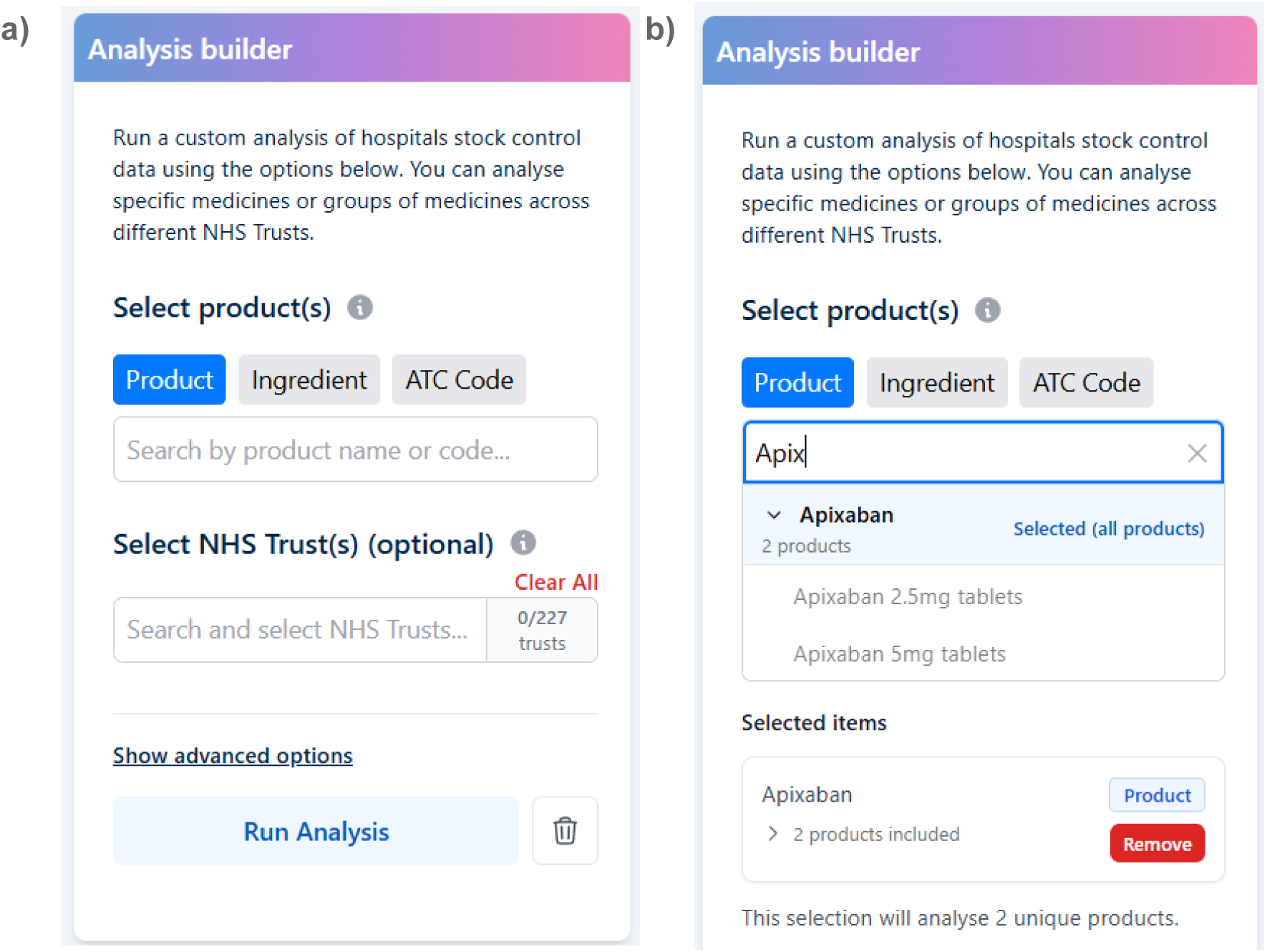
*Analysis builder* for custom analyses on OpenPrescribing Hospitals. The builder supports: free text search and selection of products by the product name or identifier, ingredient name or ATC name or code **b)** free text search and selection of NHS trusts to analyse (optional).

The results for a custom analysis are returned in three sections. The first section indicates the products included in the analysis (**Figure 4a)** with basic product information including: the product name, the group the product belongs to, the ingredients within a product, and the unit of measure for the product in the quantity type chosen for the analysis. Additionally, the products included can be edited to refine an analysis.

**Figure 4.**
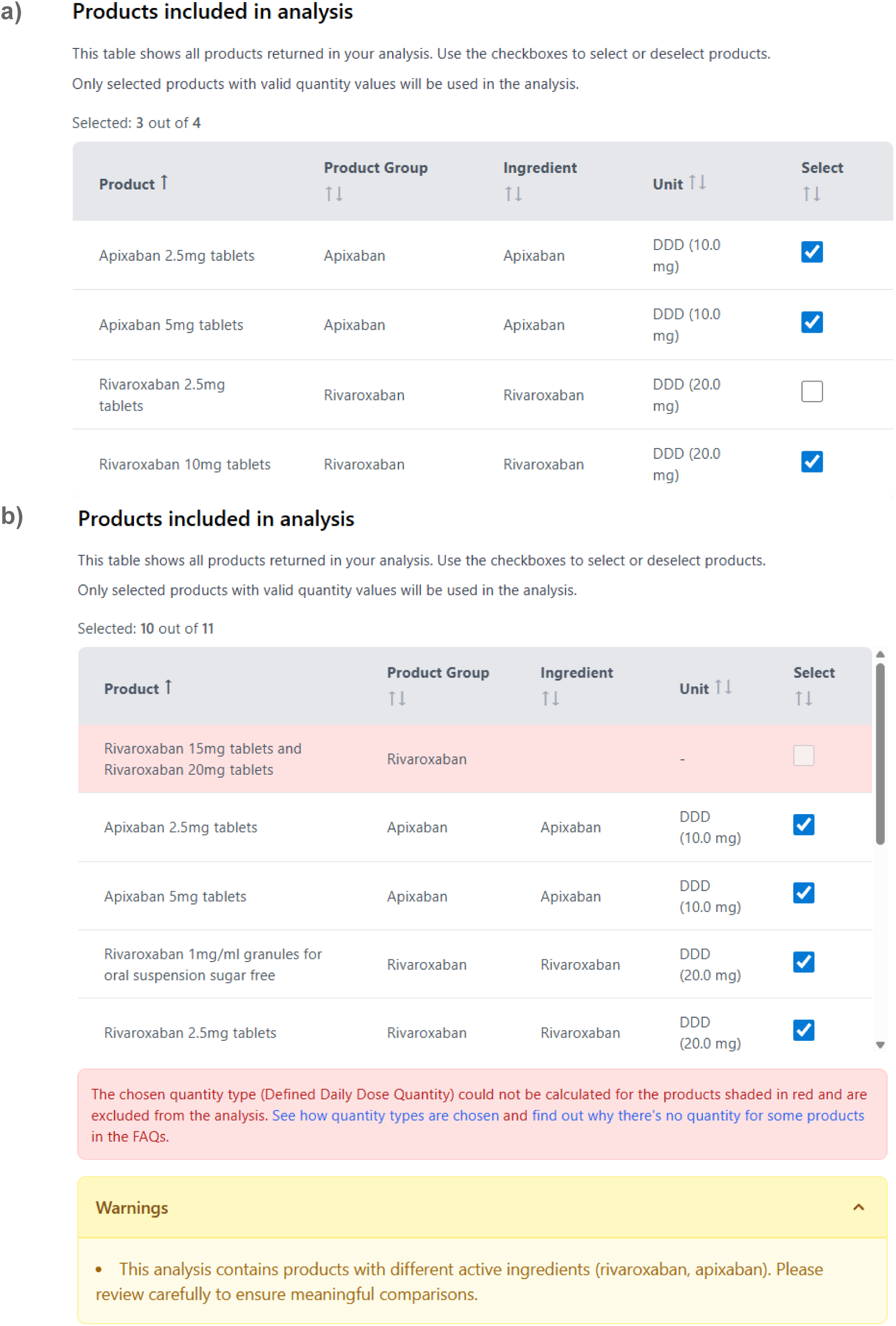
Results from a custom analysis on OpenPrescribing Hospitals **a)** The products included in an analysis, which can be edited to refine an analysis. **b)** Products are excluded and highlighted if they do not have quantity data for the quantity type used in the analysis. Warnings are also provided, where quantity aggregation for the selected products may be inappropriate.

Quantity data for the selected quantity type may not always be available for all selected products (e.g. if the quantity type is DDD quantity, but the product does not have an associated DDD value). In these cases, products are excluded from the analysis and highlighted in this table (**Figure 4b)**. Where quantity data for the chosen quantity type is available, comparison of usage across the selected products may not always be appropriate. Warnings are provided alongside this table in the following circumstances:

● The products have different units of measure for the selected quantity type. For example, analyses using *Ingredient quantity*, where some products are reported in *mls* and others are reported in *grams*.
● The selected products contain a mix of ingredients.

The second section of the analysis results displays the quantity issued for the selected product(s) as a time series chart (**Figure 5)**. Multiple chart modes are available depending on the analysis specification (**Box 3**). **Figure 5a** shows *NHS Trust* mode, with a selection of NHS trusts. **Figure 5b** shows the same product and trusts selection in *Product* mode. Examples of all of the available modes are available in **Figure S2-9.**

**Figure 5.**
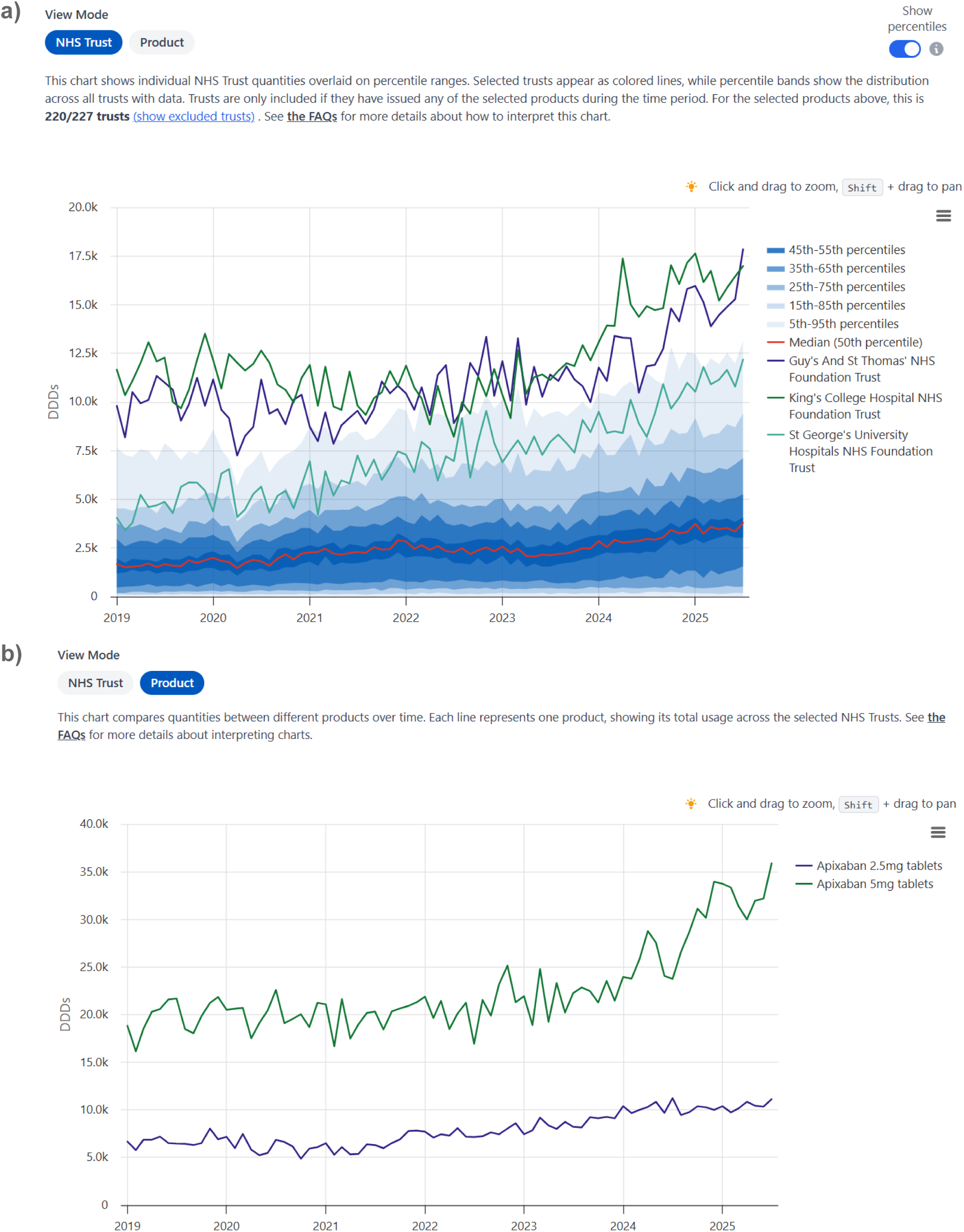
Time series charts from the results section of the *Analyse* feature on OpenPrescribing Hospitals in **a)** *NHS Trust* mode, showing the issuing patterns for selected trusts, with percentiles showing the trust-level variation nationally, **b)** *Product* mode, showing the aggregated usage for each product across the selected trusts.

**Box 3**. Chart modes available in analysis results generated from custom analyses with the Analyse feature on OpenPrescribing Hospitals.

**NHS Trust** - The default chart, available for all analyses. This mode displays percentile charts, demonstrating variation in issuing of the selected products across all NHS trusts in England issuing them. If trusts are selected in the *Analysis builder*, the data for each trust is plotted individually with the percentiles. Percentiles can be toggled to restrict this chart to only the traces for individual trusts.

**ICB mode** - The aggregated issuing of all selected products by all NHS trusts within each ICB. Only available when no trusts are selected.

**Region** - The aggregated issuing of all selected products by all NHS trusts within an NHS region. Only available when no trusts are selected.

**National** - The aggregated issuing of all selected products by all NHS trusts in England. Only available when no trusts are selected.

**Product** - The issuing of each selected product (eg apixaban 5mg tablets) aggregated across all trusts in England, if no trusts are selected, or across the selected trusts if they are. **Product group** - The issuing of the selected products aggregated by product group (e.g. all apixaban products), across all trusts in England, if no trusts are selected, or across the selected trusts if they are.

**Ingredient** - The issuing of the selected products aggregated by ingredient, across all trusts in England, if no trusts are selected, or across the selected trusts if they are.

**Unit** - The issuing of the selected products aggregated by unit of measure, across all trusts in England, if no trusts are selected, or across the selected trusts if they are.

The final section of the analysis results displays the total quantities for the selected products over the following time periods: full period of the SCMD, latest month with data, latest year, financial year to date (FYTD) (**Figure 6)**. The totals in this table are aggregated by the same modes available in the chart selection.

**Figure 6.**
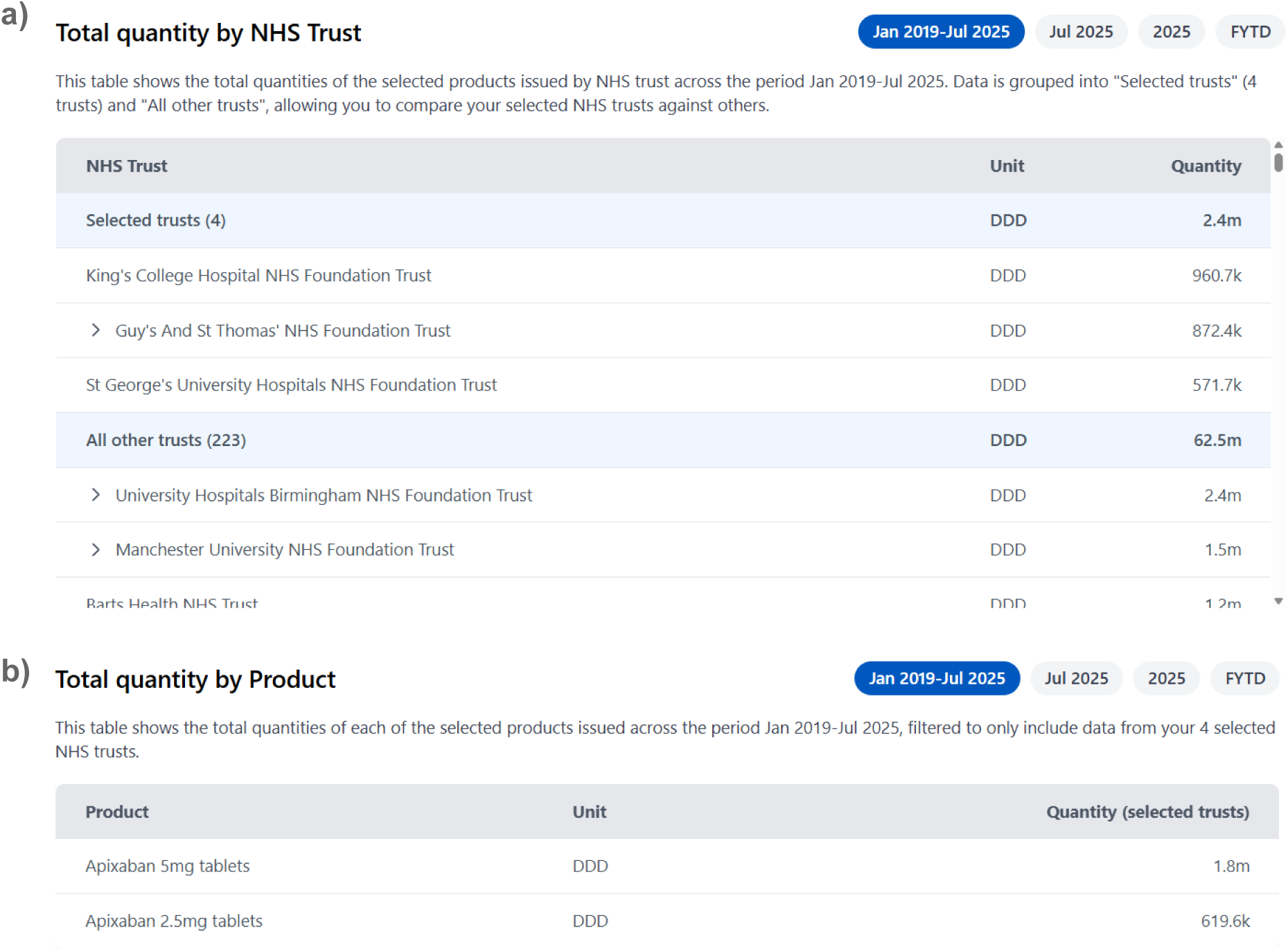
An example of the totals table in the results section of a custom analysis using the OpenPrescribing Hospitals Analyse feature. The totals table shows aggregations depending on the selected mode. **a)** The total across each trust **b)** the total for each selected product within the selected trusts.

## Results

### Use cases

Below we give example use cases for the features of OpenPrescribing Hospitals described above. These are non-exhaustive; we encourage users to explore the platform to investigate other use cases.

### Finding data quality issues

The *Submission History tool* can be used to explain unexpected results in custom analyses run using the *Analyse* feature. As an example, **Figure 7a** shows a transient drop in the number of apixaban tablets issued between February 2020 and November 2021 by an individual NHS trust. This drop aligns with a drop in the number of unique products submitted to the SCMD in the same time period, for the same trust (**Figure 7b)**. The *Submission History* tool shows that this isn’t a true change in issuing practice, but rather a data quality consideration that would be hard to identify otherwise.

**Figure 7.**
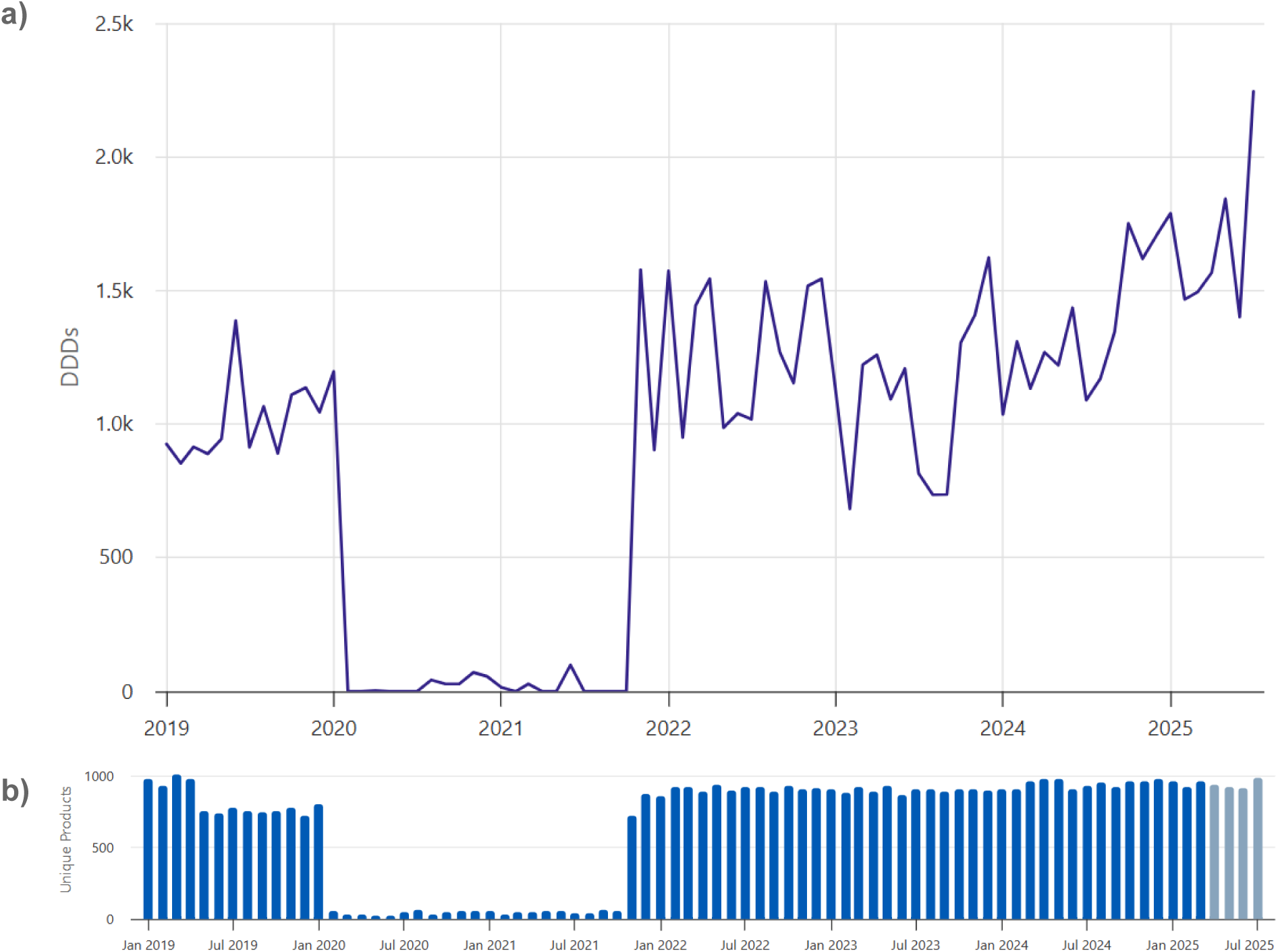
a) Results of a custom analysis of apixaban tablets in a single trust using the *Analyse* feature on OpenPrescribing Hospitals **b)** The submission history for the same trust.

### Rapid feedback on medicines use

The *Measures tool* can be used to provide rapid local feedback against national medicines standards. For example, the measure shown in **Figure 8** shows the number of lidocaine patches issued in a selection of trusts, relative to national trust-level percentiles. Lidocaine patches are included in a list of items published by NHS England which should not be prescribed in primary care due to a lack of robust evidence of clinical effectiveness^53^. In most cases, items which should not be used in primary care should not be used in local hospitals either, although some exceptions do exist.

**Figure 8.**
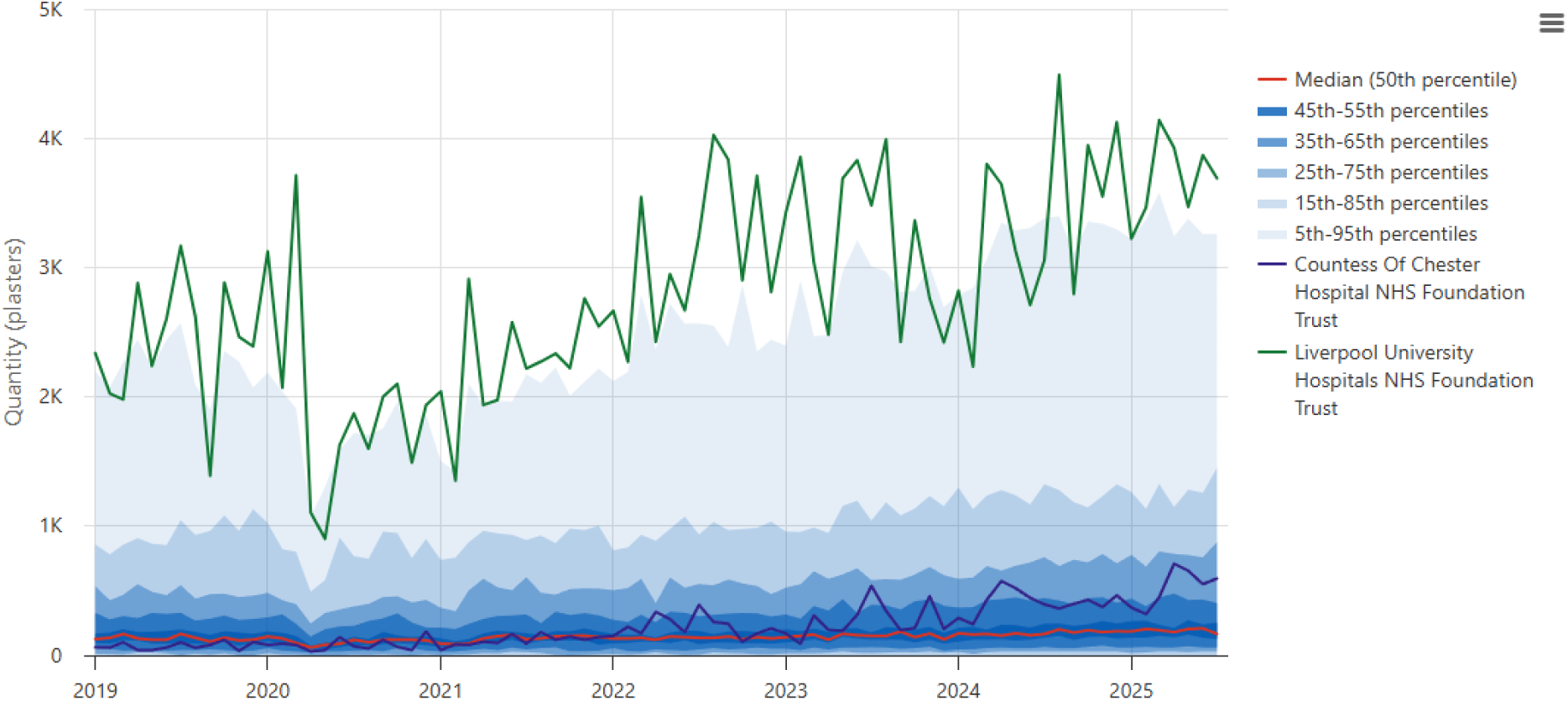
A measure on OpenPrescribing Hospitals showing the number of lidocaine patches issued each month across NHS trusts in England. This shows the trend for two selected individual NHS Trusts, relative to percentiles calculated across all NHS trusts in England.

A full list of the m*easures* available on OpenPrescribing Hospitals as of October 2025 is shown in **Table S1**.

### Monitoring innovation

The *Analyse* feature can be used to monitor the extent and variation of uptake of new medicines. For example, in May 2021 andexanet alfa was recommended by the National Institute for Health and Clinical Excellence as an option for managing major gastrointestinal bleeding in patients taking the anticoagulants apixaban or rivaroxaban^54^. Andexanet alfa is expensive and its risk-benefit profile is not well established^55,56^. **Figure 9** shows the results of a custom analysis of andexanet alfa issuing in a selection of NHS trusts using the OpenPrescribing Hospitals *Analyse* tool. It highlights wide variation in the speed and volume of uptake across trusts. This may be due to variation in local protocols^57^, which are influenced by individual clinicians’ interpretation of the evidence, raising important questions for policy makers and those responsible for local clinical guidance implementation.

**Figure 9.**
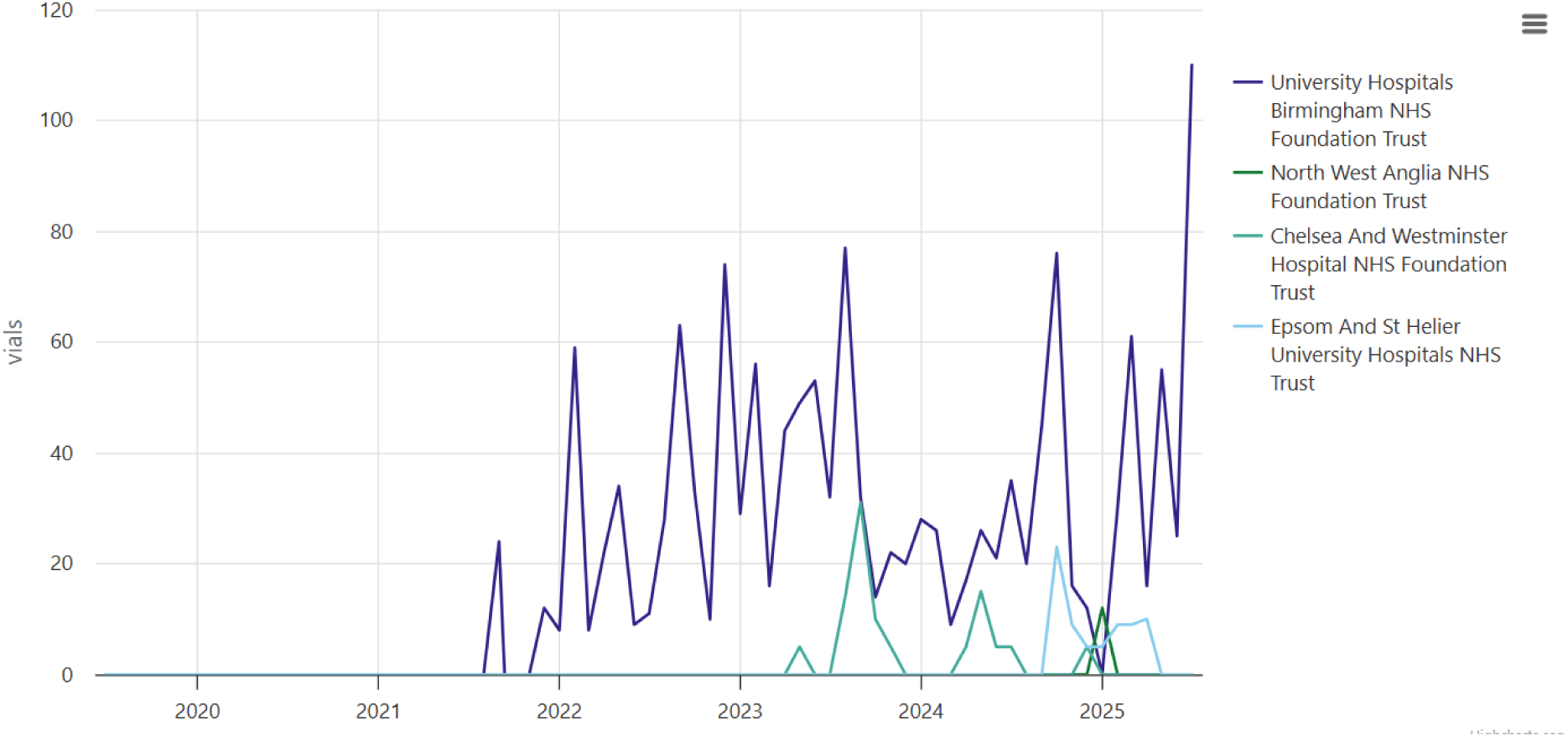
Results of a custom analysis run using the *Analyse* feature, showing the number of vials of andexanet alfa issued across all NHS trusts in England between January 2019 and July 2025.

## Discussion

OpenPrescribing Hospitals is the first openly available platform allowing *comprehensive* analysis of hospital medicines data in England. For the first time, any interested party can compare the quantity of a medicine or group of medicines issued at any trust, with aggregations of trusts within ICBs or NHS regions in England.

### Strength and weaknesses

The SCMD is a rich data source with potential to inform safer and better value care in the NHS. It is a welcome addition to the open data hosted by the NHS BSA. However, its usability is limited by the technical skill required to analyse the data. The OpenPrescribing Hospital platform provides a user-friendly interface to the SCMD. It enables anyone inside and outside the NHS *to* efficiently monitor medicines used in secondary care. For the first time, *any interested party* can monitor national initiatives^51^ for hospital trusts in their area including: low value medicines prescribing^53,58^; environmentally friendly inhalers^59^; and uptake of new treatments^60^. Prior to OpenPrescribing Hospitals, such analyses were time-consuming and limited to those, predominantly within the NHS, with password protected access to available analytic tools ^4,19,21^.

Beyond open access to the platform, another strength of this work is the transparency of the implementation. All of the source code for the data preparation and the platform are openly available^45^. We have produced extensive documentation on the challenges of working with the SCMD and how we have approached processing and presenting the data within the OpenPrescribing Hospitals platform^46,61^.

We acknowledge some limitations with the initial version of the platform. There are four major limitations inherent with the primary data source, the SCMD. Firstly, the SCMD reports stock control data, rather than individual patient prescriptions and therefore cannot differentiate when products may have been issued but not administered to a patient. Secondly, the data within the SCMD is available based on the generic name of the medicines. Brand level information is not available. As a result, we are unable to report the use of biosimilar medications which is an important national cost saving medicines priority ^62–64^ as well as any other prescribing measures that require brand level information. As an example, immunosuppressants such as tacrolimus, used in organ transplant, have a narrow therapeutic window and should be prescribed by brand to reduce the chance of organ transplantation rejection ^26,65,66^. Third, cost is reported in the SCMD as indicative cost. This does not reflect the actual cost paid by hospitals as each hospital can have different purchasing prices based on confidential price agreements. Consequently, it has limited utility. Finally, there are known data quality issues that have the potential to provide misleading results on OpenPrescribing Hospitals. Some of these are described above and are mitigated as part of the platform, but there are likely further data quality issues we have not described. Despite these limitations we believe the current version of SCMD can generate transformative insights on quality and safety of medicines issued in hospitals.

Unlike primary care, which has well defined catchment populations, it is difficult to determine the relative populations secondary care organisations serve. As a result, care must be taken when analysing the quantities of products issued across multiple trusts; variation does not necessarily represent variation in clinical practice, but could instead be indicative of the varying size and clinical specialisation of the trusts. We do not attempt to identify suitable denominator populations in custom analyses. We do however, support this in *Measures*, which by design, include denominators that allow for normalisation. We will investigate and support appropriate denominators for custom analyses as the platform develops.

### Findings in context

Over the past decade, the cost of medicines issued in hospitals has nearly doubled from £5.8 billion in 2013 - 2014 to £10.3 billion in 2023 - 2024 (+77.6%)^67,68^. This is a disproportionate increase compared with the rise in the Department of Health and Social Care spending in the same period (2013/2014 £149.7 billion - 2023/2024 £195.7 billion (+23.5%) (real-term spending for 2024-2025 prices)^69^. Hospital medicines represent a large and rapidly growing share of NHS spending. Public data on how medicines are used within hospitals allows for identification of excessive spending, particularly as hospitals are where the newest and most costly treatments are most likely to be used. Beyond cost, it also allows assessment of unwarranted variation and suboptimal care. Greater scrutiny of medicines use in hospitals is needed, and an openly available and accessible platform such as OpenPrescribing Hospitals facilitates analyses of this data within the NHS and by interested third parties.

OpenPrescribing Hospitals is the first openly available tool which can be used to carry out comprehensive analyses of individual medicines or groups of medicines at individual NHS hospital trusts in England. It was influenced by the Hospital Medicines Usage Data Explorer, an open-source tool for basic exploration of the SCMD^18^, which through open working methods supported knowledge sharing and more rapid development of the OpenPrescribing Hospitals platform^70^.

There are other platforms available for the analysis of hospital medicines ^19–22^. These platforms are available for those with password protected access which is predominantly those working within the NHS. All measure stock issued rather than individual patient level prescribing. The most similar platform is the DEFINE tool^19^, which has data on actual cost and medicines issued at brand level and is used by those with access, in the NHS. Detailed comparison with OpenPrescribing Hospitals is challenging due to the lack of openly available implementation details for these platforms^70^.

### Policy implications

National collection of GP prescribing data has taken place since the late 1980s^71,72^, with restricted access originally limited to selected NHS staff. From September 2011, however, prescribing data from general practice has been released every month as openly available data, with detail down to the level of individual practices and without information governance restrictions. This shift has enabled the development of a broad landscape of analyses, digital tools, and local support services. Medicines optimisation teams within NHS commissioning organisations now routinely use these data to review trends and provide feedback to practices, while online platforms make prescribing patterns accessible to the wider public. Researchers have also used the dataset to explore a wide variety of questions, and evidence shows that structured feedback to GPs based on these data can lead to measurable improvements in prescribing including substantial savings ^25–28^.

The NHS has now released similar data for hospital medicines data and we have built the first comprehensive widely available tool to integrate the data. We recommend three things that need to be done.

1. Funding bodies should offer funding streams to support data infrastructure projects that enable better use of data to improve patient care^70,73,74^. Funding to develop data infrastructure as a shared resource is difficult to acquire. Traditional academic funding routes do not typically support data infrastructure products or the ongoing maintenance costs of running a platform such as OpenPrescribing Hospitals. This platform has been developed on a modest budget through funding by the NHS England Primary Care and Medicines Analytics Unit.
2. The NHS should support dissemination of this openly available tool, as a complementary tool to those available in private to a small number of NHS experts. In primary care, the openly available OpenPrescribing platform has 20,000 unique users per month and has had a demonstrable impact on prescribing in primary care ^25,26^. The OpenPrescribing Hospitals platform was launched in March 2025. As of October 2025, the platform has had over 4,500 unique visitors and has supported over 5,500 custom analyses.
3. Medicines Optimisation teams in primary care are highly skilled in analysing GP prescribing data, which they use to scrutinise medicines use within their organisations and to implement changes in clinical practice. The same skills are needed across Integrated Care Systems to assess value and effectively deliver change based on sound data and analytics. The OpenPrescribing Hospitals platform supports hospital pharmacy teams, and other organisations responsible for secondary care medicines use, to achieve this.

## Administrative

### Data and software availability

The data available through the OpenPrescribing Hospitals platform is a curated aggregation of the following data sources:

- Secondary Care Medicines Dataset, openly available at https://opendata.nhsbsa.net/dataset/finalised-secondary-care-medicines-data-scmd-with-indicative-price
- Organisation Reference Data, openly available at https://digital.nhs.uk/developer/api-catalogue/organisation-data-service-ord
- NHS dictionary of medicines and devices (dm+d), openly available at https://www.nhsbsa.nhs.uk/pharmacies-gp-practices-and-appliance-contractors/nhs-d ictionary-medicines-and-devices-dmd
- Anatomical Therapeutic Chemical/Defined Daily Dose (ATC/DDD), available at https://atcddd.fhi.no/atc_ddd_index/

The source code for the data pipeline and platform is available at <u>github.com/bennettoxford/openprescribing-hospitals</u> and is made available under MIT licence. The OpenPrescribing Hospitals platform is openly available at <u>openpresribing.net/hospitals</u>.

### Contributorship

**Conceptualization**: Louis Fisher, Christopher Woods, Richard Croker, Stephen Black, Sebastian Bacon, Ben Goldacre, Brian MacKenna, and Victoria Speed.

**Data curation**: Louis Fisher, Christopher Woods, Stephen Black, and Victoria Speed. **Formal analysis**: Louis Fisher, Christopher Woods, Stephen Black, and Victoria Speed. **Funding acquisition**: Ben Goldacre and Brian MacKenna.

**Investigation**: Louis Fisher, Christopher Woods, Stephen Black, Brian MacKenna, and Victoria Speed.

**Methodology**: Louis Fisher, Christopher Woods, Richard Croker, Stephen Black, Brian MacKenna, and Victoria Speed.

**Project administration**: Louis Fisher, Christopher Woods, Brian MacKenna, and Victoria Speed.

**Resources**: Sebastian Bacon, Ben Goldacre, and Brian MacKenna.

**Software**: Louis Fisher, Christopher Woods, and Victoria Speed. **Supervision**: Ben Goldacre, Brian MacKenna, and Victoria Speed. **Validation**: Louis Fisher, Christopher Woods, and Victoria Speed.

**Visualisation**: Louis Fisher, Christopher Woods, Stephen Black, Brian MacKenna, and Victoria Speed.

**Writing - original draft**: Louis Fisher and Victoria Speed.

**Writing - review & editing**: Louis Fisher, Christopher Woods, Andrew Brown, Richard Croker, Stephen Black, Helen J. Curtis, Rose Higgins, Arina Tamborska, Sebastian Bacon, Ben Goldacre, Brian MacKenna, and Victoria Speed.

### Guarantor

The guarantor for this work is Victoria Speed.

### Ethical approval

This study uses open, publicly available data, therefore no ethical approval was required.

