## Supplementary material for "OpenPrescribing Hospitals: An open access analytics platform for hospital medicines use in England"

**Table S1.** Measures available on the OpenPrescribing Hospitals platform (October 2025).

| Title | Tags | What is it? | How is it calculated? | Why does it matter? |
|---|---|---|---|---|
| Discontinuation of insulin detemir | Safety | Total count of insulin detemir pre-filled devices / cartridges issued. | We display the total number of insulin detemir pre-filled devices / cartridges issued. | Novo Nordisk are discontinuing all formulations of Levemir®, the only insulin detemir product available in the UK, with supply due to exhaust by December 2026.<br><br>The Primary Care Diabetes & Obesity Society (PCDOS) and Association of British Clinical Diabetologists (ABCD) have written guidance to support clinicians in selecting and safely initiating alternative basal insulins. The guidance recommends that local teams diversify prescribing across the available options to reduce supply risks. Given the significant regional variation in prescribing, it also advises using data to support planning.<br><br>This OpenPrescribing Hospitals measure, alongside its primary care OpenPrescribing counterpart provides data on variation across primary and secondary care to support this planning. |
| Uptake of atezolizumab subcutaneous | Efficiency | The percentage of atezolizumab that is issued as the subcutaneous formulation. | We divide the number of products issued of the atezolizumab subcutaneous formulation by the total number of products issued of atezolizumab. We then multiply that by 100 to obtain the % each month. | Atezolizumab can be used in the treatment of a range of common cancers. If given intravenously the infusions can take 30-60 minutes. This uses a lot of nursing time and most importantly the patients time. In August 2023 a new formulation of atezolizumab designed to be administered as a subcutaneous injection in a fraction of the time received approval for use in the UK. NHS England announced in August 2023 that it would be supporting the introduction of subcutaneous atezolizumab. Although the majority of patients starting treatment with atezolizumab are expected to switch to the new subcutaneous injection, there may be a small number who remain on the intravenous infusion. |
| Uptake of nivolumab subcutaneous | Efficiency | The percentage of nivolumab that is issued as the subcutaneous formulation. | We divide the number of products issued of the nivolumab subcutaneous formulation by the total number of products issued of nivolumab. We then multiply that by 100 to obtain the % each month. | Nivolumab can be used in the treatment of a range of common cancers. If given intravenously the infusions can take 30-60 minutes. This uses a lot of nursing time and most importantly the patients time. In April 2025 a new formulation of nivolumab designed to be administered as a subcutaneous injection in a fraction of the time received approval for use in the UK. NHS England announced in April 2025 that it would be supporting the introduction of subcutaneous nivolumab. Although not all patients will be eligible, around two in five of those currently receiving intravenous nivolumab are expected to switch to the new subcutaneous injection and most eligible new patients are also expected to begin on the injectable form of nivolumab. |
| Proportion of sodium-glucose co-transporter 2 (SGLT-2) inhibitors not prescribed as dapagliflozin tablets | Low value prescribing | The proportion of sodium-glucose co-transporter 2 (SGLT-2) inhibitors (excluding combination products) not prescribed as dapagliflozin tablets. Combination products are excluded as they do not have | We identify all SGLT-2 inhibitor products using Virtual Therapeutic Moieties (VTM) codes. The quantity of each product issued as DDDs is calculated. We then divide the DDDs of SGLT-2 inhibitors excluding dapagliflozin by the total DDDs of all SGLT-2 inhibitors and then multiply that by 100 to obtain a | Dapagliflozin is a sodium-glucose co-transporter 2 (SGLT-2) inhibitor commonly prescribed in type 2 diabetes, the treatment of chronic heart failure, and chronic kidney disease.<br><br>Dapagliflozin lost its patent in August 2025. From September 2025 the NHS tariff price is likely to fall, as it will be based on the generic price, rather than the branded product. NICE guidance on the treatment of diabetes states that if 2 drugs in the same class are appropriate, choose the option with the lowest acquisition cost. Dapagliflozin is likely to have cost-saving opportunities when compared with other SGLT-2s.<br><br>**Note**: Dapagliflozin was licensed for the treatment of chronic heart failure with reduced ejection fraction] in February 2021, and later for chronic kidney disease in July 2025. |

| | | | | |
|---|---|---|---|---|
| | | specified Defined Daily Doses (DDDs) and the quantity of SGLT-2 inhibitors issued as combinations is low.<br><br>There is a similar measure on OpenPrescribing for primary care, which reports the use of generic dapagliflozin. In OpenPrescribing Hospitals, branded and generic products cannot be distinguished. For this reason, we report dapagliflozin use as a proportion of all SGLT-2 inhibitor use. | percentage each month. | Empagliflozin was also licensed for the treatment of chronic heart failure with reduced ejection fraction in March 2022 and for chronic kidney disease in December 2023.<br>Increasing use of dapagliflozin (and therefore reduction in the proportion of SGLT-2 inhibitors not prescribed as dapagliflozin) is the likely reason for the pattern seen in this measure prior to September 2025. |
| Unlicensed omeprazole liquid | Safety | The proportion of all omeprazole liquid issued as unlicensed products. | We divide the number of Defined Daily Doses (DDDs) of unlicensed liquid omeprazole products by the total number of DDDs of all liquid omeprazole products and then multiply by 100 to obtain a percentage each month. | An unlicensed medicine (sometimes called a special or special-order medicine) is a medicine that does not hold a UK marketing authorisation (product licence) from the Medicines and Healthcare products Regulatory Agency (MHRA). That means it has not gone through the usual MHRA approval process for safety, quality, and efficacy for its intended use.<br><br>The MHRA states that prescribers should be satisfied that an alternative, licensed medicine would not meet the patient's needs before prescribing an unlicensed medicine see, MHRA Off-label or unlicensed use of medicines: prescribers' responsibilities.<br><br>Licensed formulations of omeprazole liquid began to become available in 2020, and prescribers should consider using these in preference to unlicensed products. |
| Low value prescribing - lidocaine plasters | Value, Low value prescribing | Total lidocaine plasters issued | We display the total number of lidocaine plasters issued | NHS England guidance identifies a number of items which provide low value when prescribed. This guidance has mainly been aimed at primary care but we think it is important to also monitor a selection of these in hospitals - read more about why in our blog.<br><br>NHS England guidance states:<br><br>*Lidocaine plasters are licensed for symptomatic relief of neuropathic pain associated with previous herpes zoster infection (PHN) in adults.*<br><br>*NICE guidance on chronic pain does not recommend lidocaine plasters for treating neuropathic pain.*<br><br>*The Specialist Pharmacy Service carried out an evidence review on behalf of NHS England to inform* |

| | | | | |
|---|---|---|---|---|
| | | | | *the joint clinical working group's recommendations. Based on this review, lidocaine plasters can be prescribed only for patients who are intolerant of first-line systemic therapies for PHN or where these therapies have been ineffective.*<br><br>*Lidocaine plasters are not an alternative to an opioid-based medicine when concerned about dependence and withdrawal.*<br><br>NHS England recommend that GPs:<br>- Do not initiate.<br>- Deprescribe in patients currently prescribed this medicine.<br>- Prescribe only if no other item or intervention is clinically appropriate.<br>- Prescribe only if no other item or intervention is available.<br>- Prescribe only if for a named indication in this guidance.<br><br>For guidance on when prescribing may be appropriate in some exceptional circumstances, please see the full NHS England guidance document. |
| Environmentally friendly inhalers | Greener NHS | Percentage of inhaler doses that are from metered dose inhalers (MDIs), excluding salbutamol inhalers. | We divide the number of MDI inhaler doses by the total number of inhaler doses and then multiply that by 100 to obtain the % each month. We do not include salbutamol inhalers in these calculations or inhalers which are used outside of the management of common respiratory conditions such as asthma or chronic obstructive pulmonary disorder. | The updated BTS/NICE/SIGN guidance on asthma management recommends basing inhaler choice on:<br>- an assessment of correct technique<br>- the preference of the person receiving the treatment<br>- the lowest environmental impact among suitable devices<br>- the presence of an integral dose counter<br><br>The NHS has committed to reducing its carbon footprint by 80% by 2028 to 2032, including a shift to lower carbon inhalers. Dry powder inhalers (DPIs) and other newer types of inhalers like soft mist inhalers are less harmful to the environment than traditional metered dose inhalers (MDIs).<br><br>The NHS England National Medicines Optimisation Opportunities for 2024/25 identify reducing carbon emissions from inhalers as an area for improvement.<br><br>NICE has produced an inhaler decision aid to facilitate discussion about inhaler options. |
| Uptake of Phesgo® | Efficiency | The percentage of pertuzumab that is issued as the combined preparation, Phesgo®. | We divide the number of products containing trastuzumab and pertuzumab (Phesgo®) by the total number of products containing pertuzumab including Phesgo®. We then multiply that by 100 to obtain the % each month. | Trastuzumab and pertuzumab are used to treat breast cancer. If given separately, they are administered via two intravenous infusions which can take up to two and a half hours. This uses a lot of nursing time and most importantly the patients time. Phesgo® is a combination of trastuzumab and pertuzumab and is administered as a subcutaneous injection, in a fraction of the time. Phesgo® was made available in 2021 and was highlighted by NHSE as a medicines optimisation opportunity to improve equitable adoption of the most clinically and cost-effective medicines. |
| Direct Oral Anticoagulants (DOACs) not prescribed as | Value | Percentage of DOACs that are not rivaroxaban or apixaban tablets. | We divide the combined number of defined daily doses (DDDs) of DOACs that are **NOT** rivaroxaban or | In September 2024 the NHS in England released updated commissioning recommendations for DOACs, which state:<br><br>*For patients commencing treatment for atrial fibrillation (AF): subject to the criteria specified in the* |

| | | | | |
|---|---|---|---|---|
| rivaroxaban or apixaban tablets | | | apixaban tablets by the total number of DDDs of all DOACs and then multiply that by 100 to obtain a percentage each month. Rivaroxaban 15mg/20mg treatment initiation packs have been excluded from the measure as it is not possible to allocate a defined daily dose for comparison. | *relevant NICE technology appraisal guidance, clinicians should use the best value DOAC that is clinically appropriate for the patient. Apixaban and rivaroxaban (prescribed generically) are currently the joint best value DOACs.*<br><br>The NHS England National Medicines Optimisation Opportunities for 2024/25 also identify using best value direct-acting oral anticoagulants as an area for improvement.<br><br>**Please note:** OpenPrescribing Hospitals measures have a consistent way of visualising measures, where 'lower is better' and therefore in this case we show the proportion of DOACs which are NOT prescribed as rivaroxaban or apixaban tablets. There are also other oral formulations of DOACs recently introduced onto the market. We do not consider these to be as cost-effective, and they are not considered as first-line in this measure. You can read more about these details and the timeline of the commissioning recommendations in our blog. |
| Methotrexate tablets | Safety | Percentage of methotrexate tablets which are 10mg | We divide the number of methotrexate 10mg tablets by the total number of methotrexate tablets and then multiply that by 100 to obtain the % each month. | Overdose of methotrexate for non-cancer treatment was described as an NHS Never Event in 2018. To reduce the risk of harm, the British National Formulary (BNF) states that methotrexate should, usually, only be prescribed and dispensed as a single strength of tablet, usually 2.5 mg, to reduce the risk of harm from errors. |
| Low value prescribing - doxazosin modified release | Value, Low value prescribing | Percentage of doxazosin modified release of all doxazosin issued. | We divide the number of defined daily doses (DDDs) of doxazosin modified release tablets by the total number of DDDs of doxazosin immediate release and modified release tablets and then multiply that by 100 to obtain a percentage each month. | NHS England guidance identifies a number of items which provide low value when prescribed. This guidance has mainly been aimed at primary care but we think it is important to also monitor a selection of these in hospitals - read more about why in our blog.<br><br>NHS England guidance states:<br><br>*Doxazosin is an alpha-adrenoceptor blocking drug that can be used to treat hypertension and benign prostatic hyperplasia. There are two oral forms of the medication (immediate release and prolonged release) and both are taken once daily.*<br><br>*Prolonged-release doxazosin costs approximately six times more than doxazosin immediate release.*<br><br>*NICE guidance on hypertension recognises that doxazosin should be used in treatment but does not identify any benefits of prolonged release over immediate release.*<br><br>*NICE guidance recommends doxazosin as an option in men with moderate to severe lower urinary tract symptoms. It does not identify benefits of prolonged release over immediate release.*<br><br>*Due to the significant extra cost of prolonged-release doxazosin and the availability of once daily immediate-release doxazosin, the joint clinical working group considered prolonged-release doxazosin suitable for inclusion in this guidance.*<br><br>For guidance on when prescribing may be appropriate in some exceptional circumstances, please see the full NHS England guidance document. |
| Low value | Safety, | Total indicative cost | We display the indicative | NHS England guidance identifies a number of items which provide low value when prescribed. This |

| | | | | |
|---|---|---|---|---|
| prescribing - co-proxamol | Value, Low value prescribing | for co-proxamol tablets and oral suspension | spend for co-proxamol tablets and oral suspension. You can read more about indicative cost in our FAQs. | guidance has mainly been aimed at primary care but we think it is important to also monitor a selection of these in hospitals - read more about why in our blog.<br><br>NHS England guidance states:<br>*The Medicines and Healthcare products Regulatory Agency (MHRA) fully withdrew the painkiller co-proxamol from the UK market in 2007 due to safety concerns. All use in the UK is now on an unlicensed basis. Prescribing an unlicensed medicine should be in line with General Medical Council (GMC) guidance, which states suitably licensed alternatives need to be considered and the prescriber must be satisfied that there is sufficient evidence or experience of using the medicine to demonstrate its safety and efficacy.*<br><br>*Since 1985 advice aimed at the reduction of co-proxamol toxicity and fatal overdose has been provided, but this was not effective and resulted in withdrawal of co-proxamol by the MHRA. In 2011 MHRA reported that the withdrawal of co-proxamol from the UK had saved an estimated 300 to 400 lives each year from self-poisoning, around a fifth of which would have been accidental. Since the withdrawal, further safety concerns have been raised, resulting in co-proxamol being withdrawn in other countries.*<br><br>*Due to the significant safety concerns, the joint clinical working group considered co-proxamol suitable for inclusion in this guidance. Co-proxamol is no longer manufactured or supplied in the UK and any use on an unlicensed basis requires it to be imported for individual use, at an increasing cost to the NHS and the environment.*<br><br>NHS England recommend that GPs:<br><br>- Do not initiate.<br>- Deprescribe in patients currently prescribed this medicine. |
| Low value prescribing - perindopril arginine | Value, Low value prescribing | Percentage of perindopril arginine of all perindopril. | We divide the number of defined daily doses (DDDs) of perindopril arginine tablets by the total number of DDDs of perindopril arginine and perindopril erbumine tablets and then multiply that by 100 to obtain a percentage each month. | NHS England guidance identifies a number of items which provide low value when prescribed. This guidance has mainly been aimed at primary care but we think it is important to also monitor a selection of these in hospitals - read more about why in our blog.<br><br>NHS England guidance states:<br>*Perindopril is an ACE inhibitor used in heart failure, hypertension, diabetic nephropathy, and prophylaxis of cardiovascular events. The perindopril arginine salt version is more stable in extremes of climate than the perindopril erbumine salt, which gives it a longer shelf-life. However, perindopril arginine is significantly more expensive than perindopril erbumine and a PrescQIPP CIC review of the topic found no clinical advantage for the arginine salt.*<br><br>*NICE guidance on hypertension in adults recommends that prescribing costs are minimised.*<br><br>*Due to the significant extra costs of the arginine salt and the availability of the erbumine salt, the joint clinical working group considered perindopril arginine suitable for inclusion in this guidance.*<br><br>For guidance on when prescribing may be appropriate in some exceptional circumstances, please see the full NHS England guidance document. |

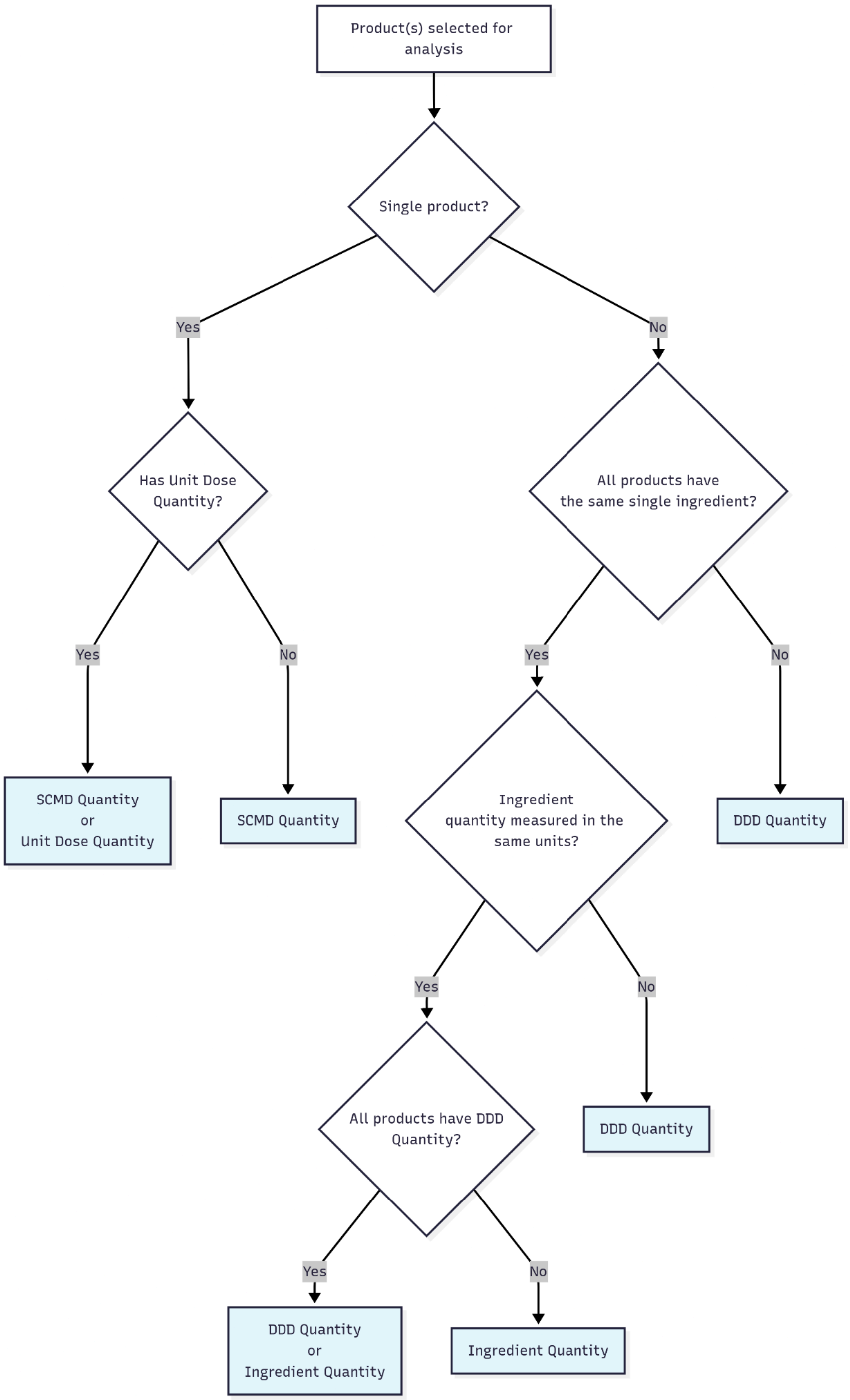


**Figure S1.** The logic used for automated selection of quantity type for custom analyses on OpenPrescribing Hospitals. This selection is designed to improve the likelihood that the quantities being compared are clinically meaningful and that quantity data is available for the greatest number of the selected products.

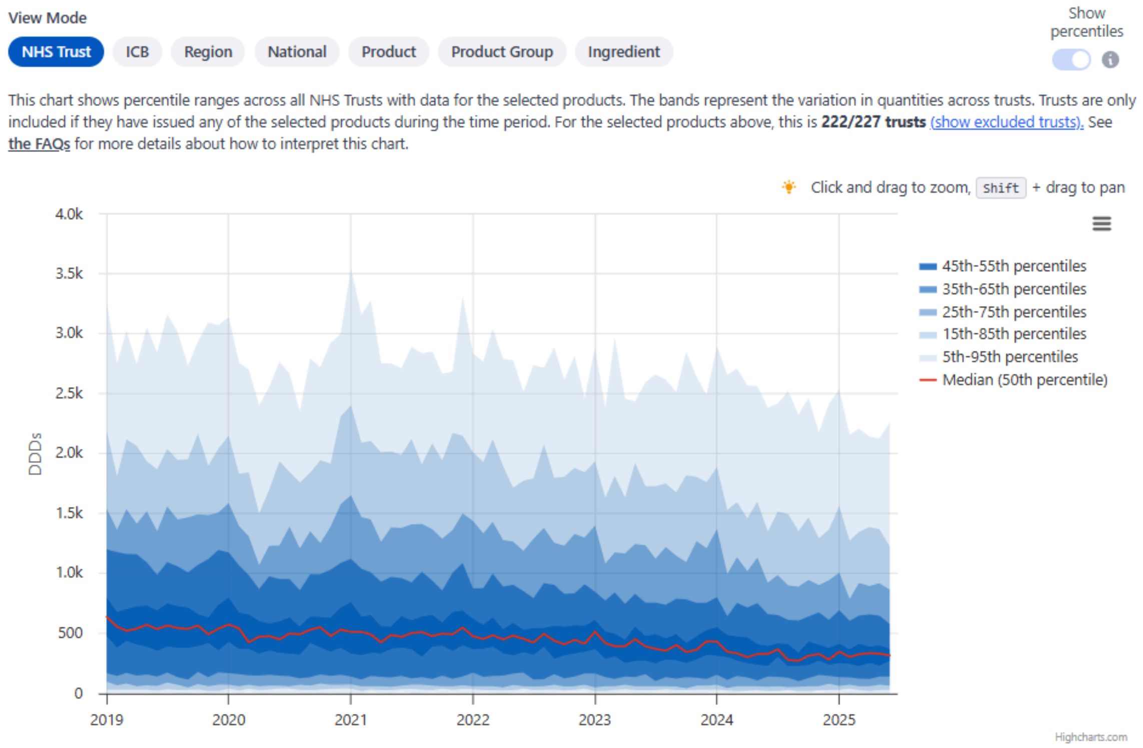


**Figure S2.** Time series charts from the results section of the Analyse feature on OpenPrescribing hospitals in NHS trust mode, with percentiles showing trust-level variation in issuing of the selected products.

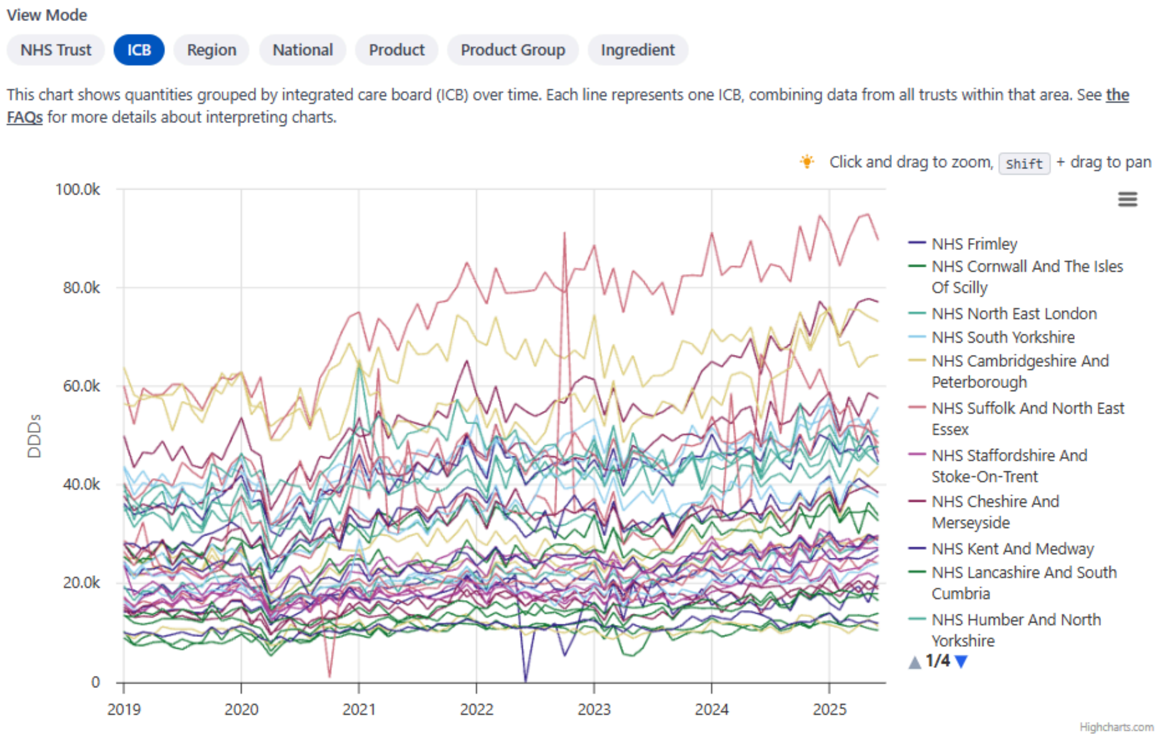


**Figure S3.** Time series charts from the results section of the Analyse feature on OpenPrescribing hospitals in ICB mode, showing the issuing quantity of the selected products in NHS trusts within each ICB.

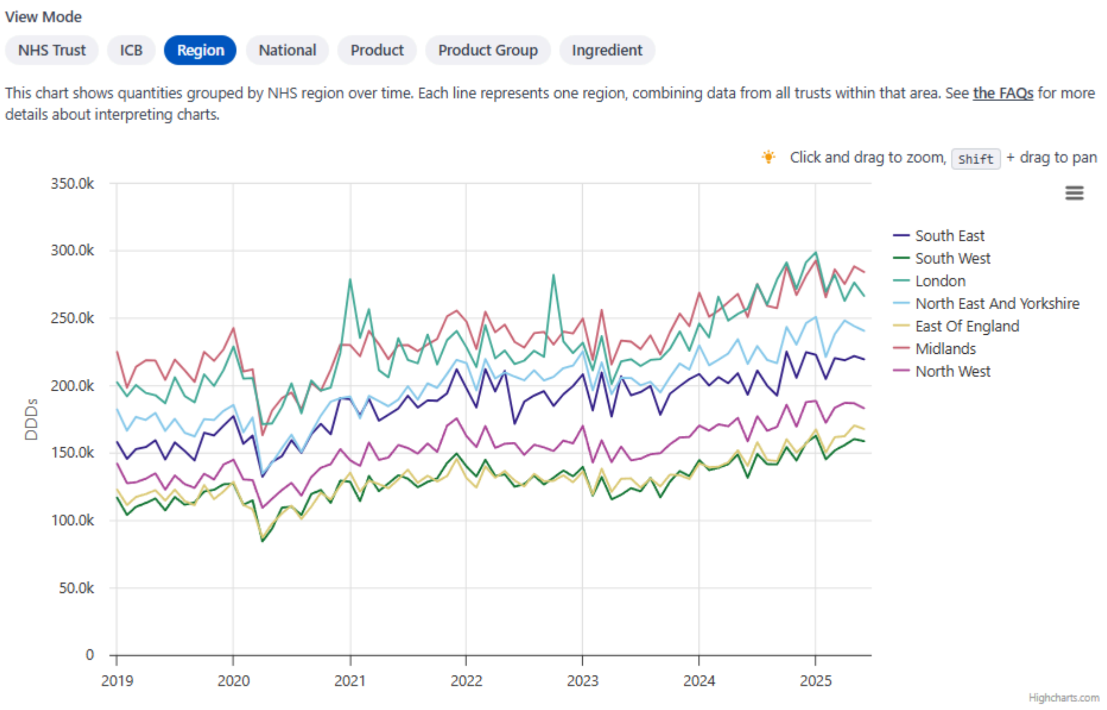


**Figure S4.** Time series charts from the results section of the Analyse feature on OpenPrescribing hospitals in region mode, showing the issuing quantity of the selected products in NHS trusts within each NHS region.

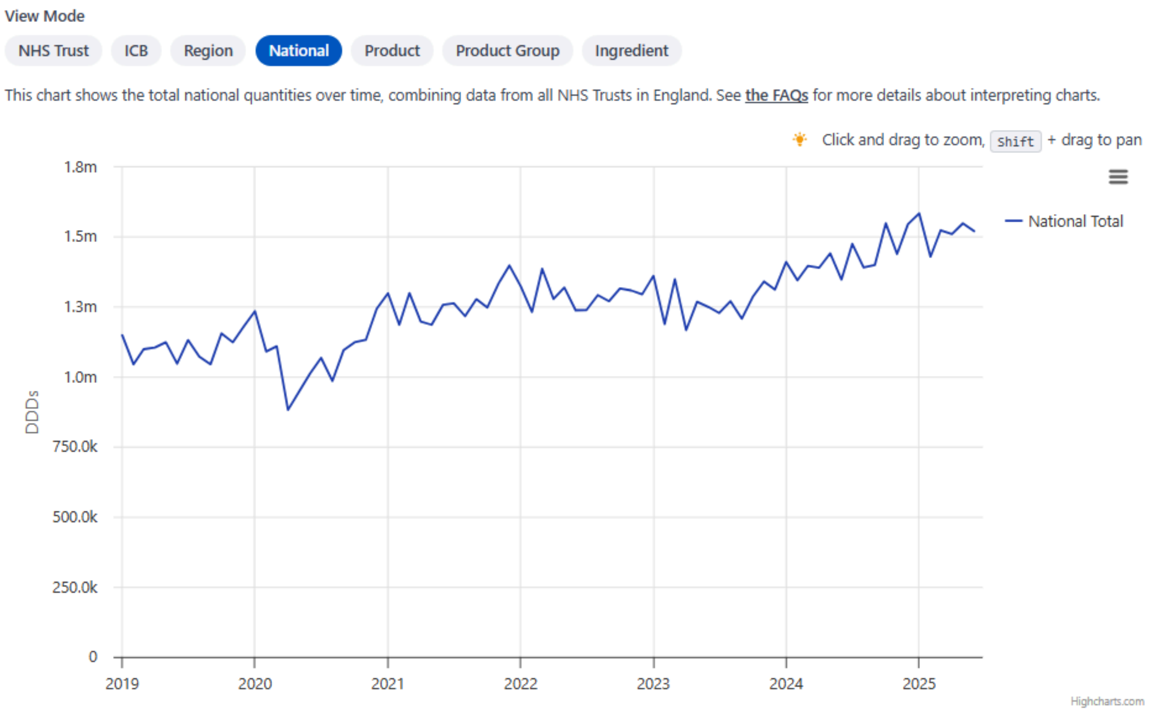


**Figure S5.** Time series charts from the results section of the Analyse feature on OpenPrescribing hospitals in national mode, showing the total issuing quantity of the selected products across all NHS trusts in England.

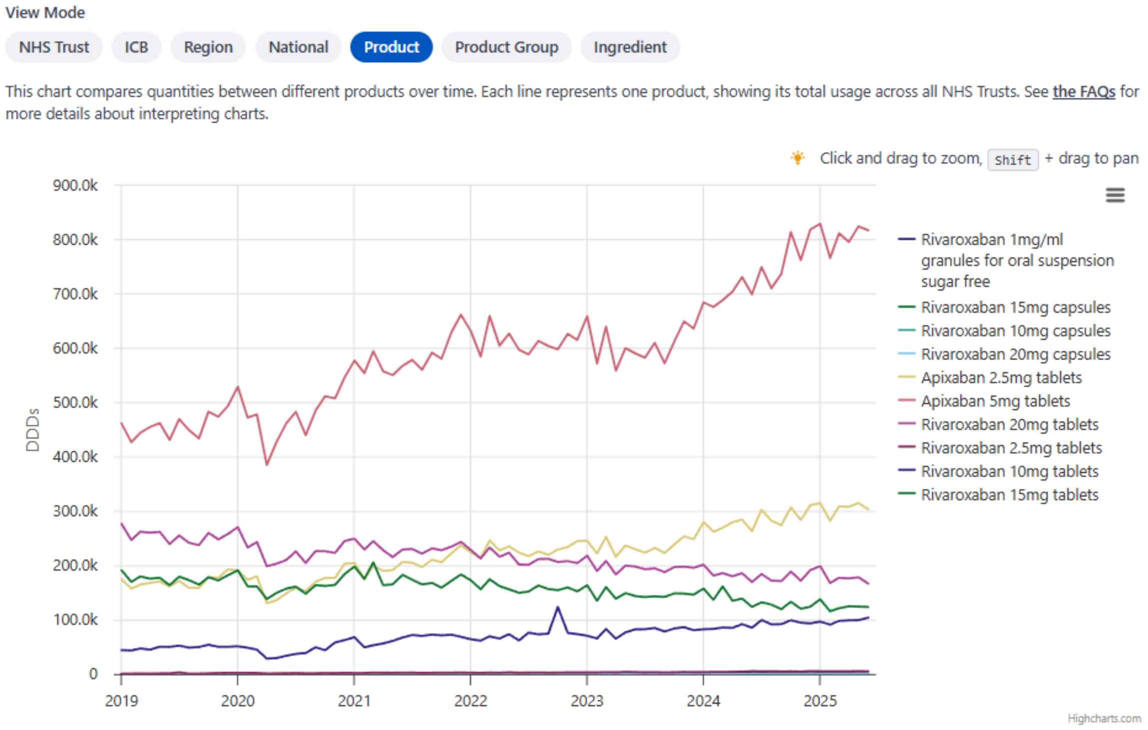


**Figure S6.** Time series charts from the results section of the Analyse feature on OpenPrescribing hospitals in product mode, showing the issuing of each selected product.

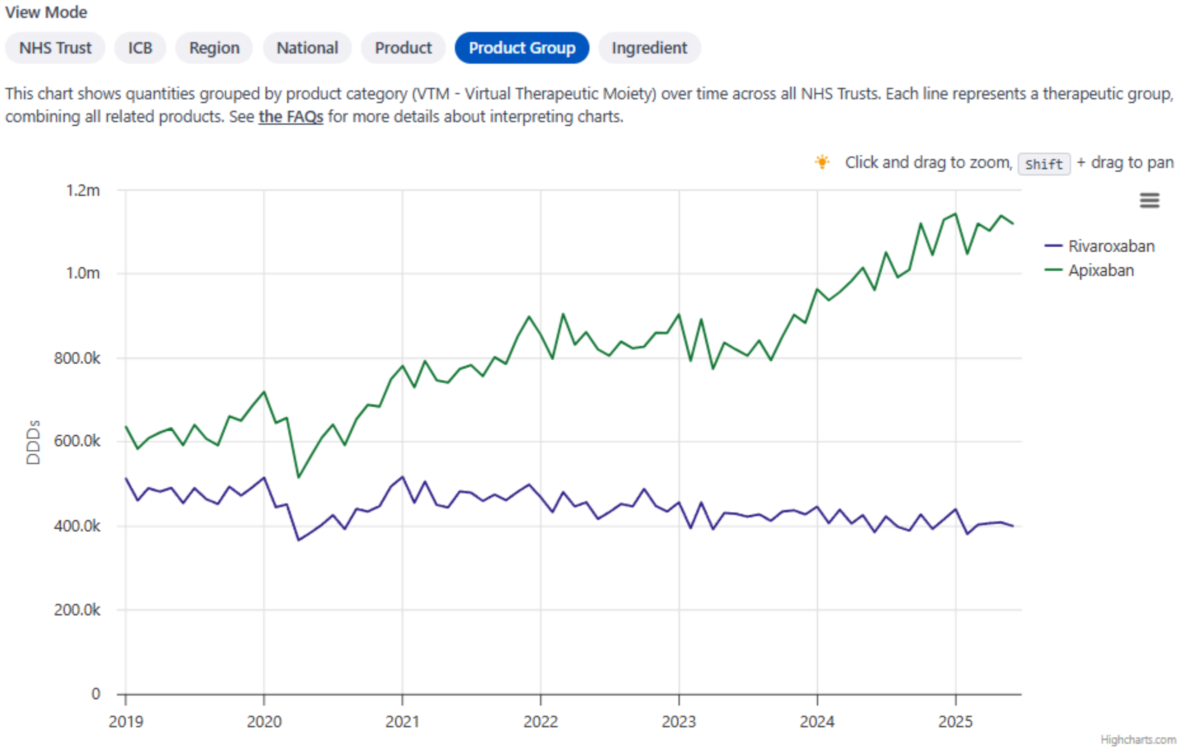


**Figure S7.** Time series charts from the results section of the Analyse feature on OpenPrescribing hospitals in product group mode, showing the issuing of products belonging to each product group (Virtual Therapeutic Moiety) represented by the selected products.

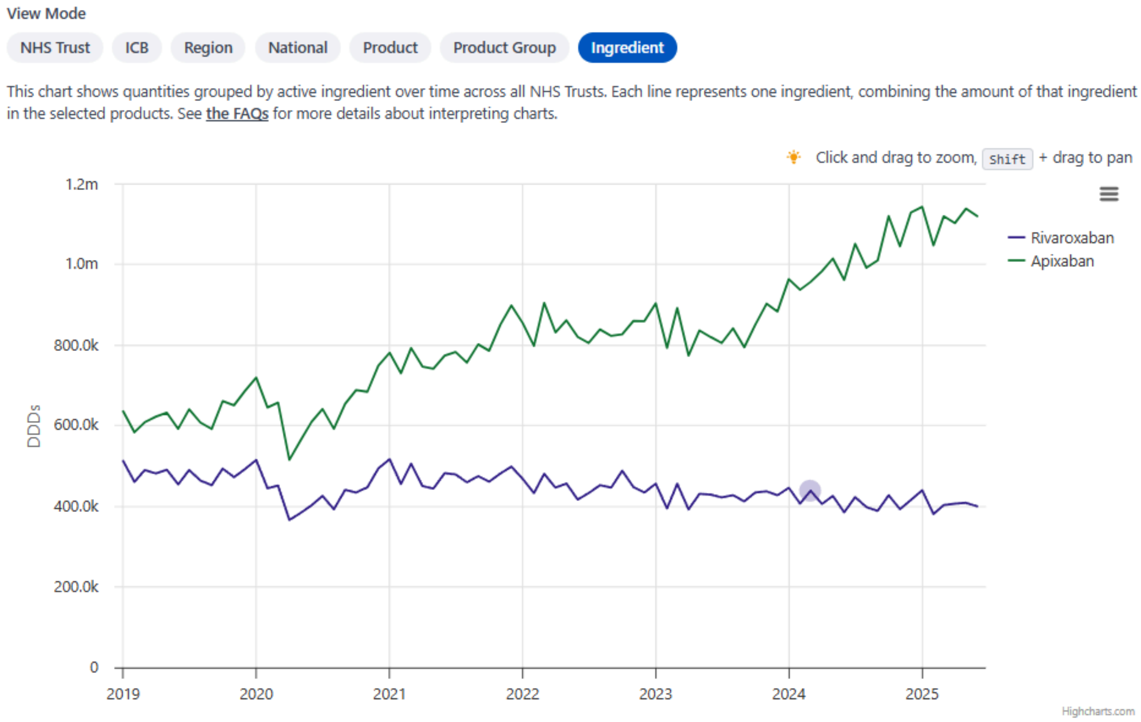


**Figure S8.** Time series charts from the results section of the Analyse feature on OpenPrescribing hospitals in ingredient mode, showing the quantity of each active ingredient within the selected products.

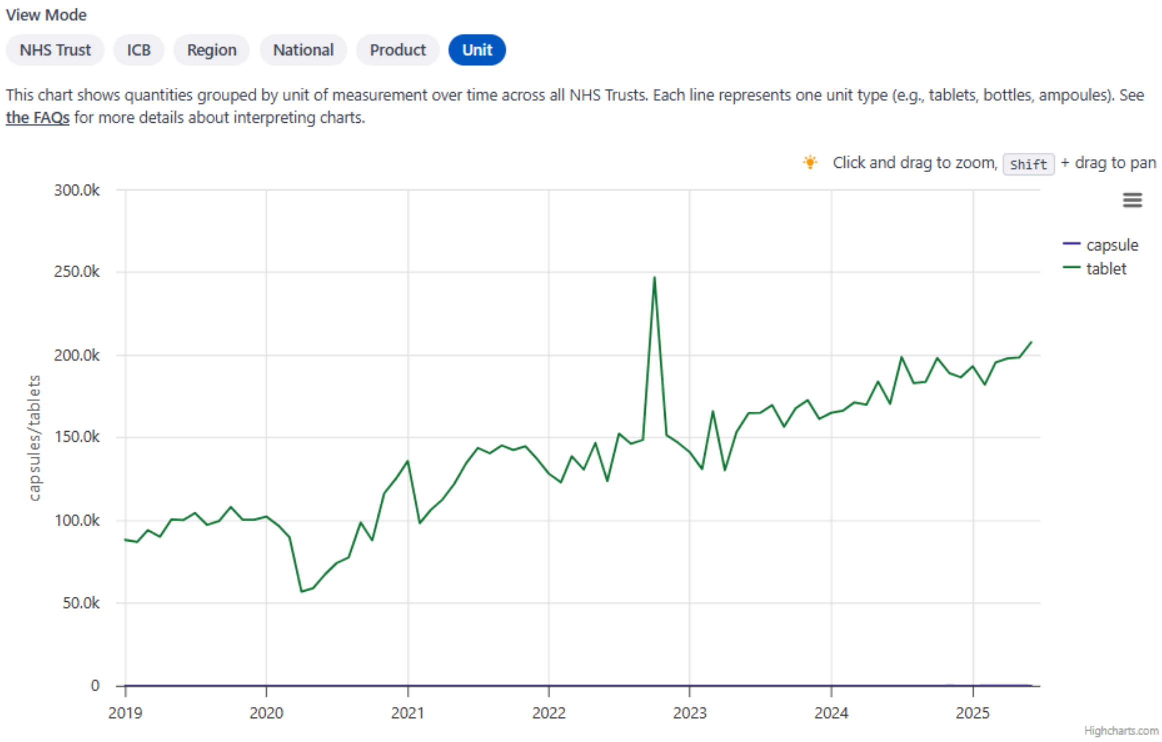


**Figure S9.** Time series charts from the results section of the Analyse feature on OpenPrescribing hospitals in unit mode, showing the quantity of the selected products broken down by unit of measure.
